# Protocol for a retrospective, comparative effectiveness study of the association between transesophageal echocardiography (TEE) monitoring used in coronary artery bypass graft (CABG) surgery and clinical outcomes

**DOI:** 10.1101/2020.05.23.20110528

**Authors:** Emily J. Mackay, Bo Zhang, Siyu Heng, Ting Ye

**Author notes:** Address for Correspondence:* Emily J. Mackay, Department of Anesthesiology and Critical Care, Perelman School of Medicine, University of Pennsylvania, Philadelphia, PA, U.S.A.

## Abstract

**Background:** Coronary artery bypass graft (CABG) surgery is the most widely performed adult cardiac surgery in the US. Transesophageal echocardiography (TEE) is an ultrasound-based cardiac imaging modality used in CABG surgery for hemodynamic monitoring and management of complications related to cardiopulmonary bypass. However, there are no comparative effectiveness studies (randomized or non-randomized) that have investigated the relationship between TEE monitoring and clinical outcomes among patients undergoing CABG surgery. Because of this lack of evidence, recommendations for TEE in CABG surgery remain indeterminate (Class II). We aim to compare the clinical outcomes of patients undergoing CABG surgery with vs without TEE monitoring. This protocol will detail how we plan to investigate the hypothesis that TEE monitoring in CABG surgery will be associated with improved clinical outcomes.

**Methods and Analysis:** This investigation will be an observational retrospective, comparative effectiveness, cohort study using Centers for Medicare and Medicaid Services (CMS) claims data from January 1, 2013 to October 15, 2015. The aim is to determine if TEE monitoring during CABG surgery is associated with improved 30-day survival, lower incidence of stroke, shorter length of hospitalization, and incidence of esophageal perforation. To alleviate the potential bias from unmeasured confounding, we propose leveraging hospitals’ (or surgeons’) preference for TEE in CABG surgery as an instrumental variable (IV). We will combine this IV technique with statistical-matching-based methods by pairing hospitals (or surgeons) with similar observed confounding variables but considerably different preference for TEE monitoring in CABG surgery. Our research design is meant to emulate a cluster-randomized encouragement experiment. The following a priori protocol will detail how we plan to execute this analysis.

## 1 Background and motivation

Coronary artery bypass graft (CABG) surgery is the most widely performed adult cardiac surgery, accounting for over half of the 300,000 cardiac surgeries performed in the US each year (The Society of Thoracic Surgeons, 2016b). While CABG surgery carries a lower risk of death (2%) (Jacobs et al., 2018), compared to that of valve surgery (5%) (Smith et al., 2011, Acker et al., 2014, Reardon et al., 2017), the decline in CABG surgery by the expansion of percutaneous coronary revascularization (Epstein et al., 2011, Riley et al., 2011, The Society of Thoracic Surgeons, 2016a) means that patients who do require CABG surgery are increasingly sicker, have a higher coronary artery disease burden, and higher rates of comorbid conditions (Aldea et al., 2009, Mokadam et al., 2011, Cao et al., 2013, Aldea et al., 2016).

Transesophageal echocardiography (TEE) is an ultrasound-based, cardiac imaging modality, frequently used in CABG surgery for hemodynamic monitoring and management of complications related to cardiopulmonary bypass (Cheung et al., 1994, Shapira et al., 2004, Charron et al., 2006, Shapira et al., 2007, Eltzschig et al., 2008, Silva et al., 2010, Hahn et al., 2013, Nishimura et al., 2014, 2017). While complications directly related to TEE are rare (Hilberath et al., 2010), unnecessary procedures performed for incidental findings diagnosed by TEE during CABG surgery has been associated with worse clinical outcomes (Krasuski et al., 2009), and guidelines for TEE monitoring in CABG surgery remain indeterminate (Hahn et al., 2013, Hillis et al., 2011). Nevertheless, because of its ability to accurately differentiate between cardiac failure and hypovolemia (Cheung et al., 1994, Charron et al., 2006, Eltzschig et al., 2008, Silva et al., 2010, Hahn et al., 2013, Nishimura et al., 2014, 2017), routine perioperative monitoring with TEE could potentially improve clinical outcomes if it led to better hemodynamic management during CABG surgery.

So far, there are no randomized comparative effectiveness studies investigating the relationship between TEE monitoring and clinical outcomes among patients undergoing isolated CABG surgery. Observational work from our group has demonstrated that the use of TEE in CABG surgery is highly variable, ranging from 11% to 91% across US states (MacKay et al., 2020). Such variability in the practice of TEE for CABG surgery presents a comparative effectiveness research opportunity to evaluate the impact of TEE use on perioperative outcomes of CABG surgery patients.

## 2 Overview of the research design

We plan to undertake an observational retrospective, comparative effectiveness, cohort study using Centers for Medicare and Medicaid (CMS) claims data from January 1st, 2013 to October 15th, 2015 to determine if TEE monitoring during CABG surgery was associated with 30-day mortality (primary outcome), length of hospitalization (secondary outcome), incidence of stroke (secondary outcome), and incidence of esophageal perforation (secondary outcome).

We anticipated that sicker patients would be more likely to receive perioperative TEE for CABG surgery. Therefore, a direct comparison between patients receiving TEE monitoring and those not receiving TEE monitoring is not warranted. Controlling for observed confounding variables helps eliminate some of the selection bias; however, bias from factors that are not measured or accounted for, e.g., patients’ ejection fraction, hemodynamics including blood pressure, cardiac index, and use of inotropic or vasopressor medications in operating room, and laboratory values, would remain.

To alleviate the potential bias from aforementioned unmeasured confounding variables, we propose to leverage hospitals’ (or surgeons’) preference for using TEE during CABG surgery as an instrumental variable. Classical approaches to preference-based-IV analyses typically rely on structural equation models or other parametrized models to control for IV-outcome confounders (Brookhart et al., 2006, Hernán and Robins, 2006). Examples include the two-stage-least-squares (2SLS) method and variants of it. Despite their convenience and simplicity, these parametric models rely on model assumptions that are often difficult to check. Statistical-matching-based methods, on the other hand, provide a tempting nonparametric alternative to structural equation models. We propose to apply a matching-based approach to the current study. In particular, we pair hospitals (or surgeons) with similar confounding variables but considerably different preference for TEE monitoring in CABG surgery. Our research design is meant to replicate a **cluster-randomized encouragement experiment** (also known as a **cluster-randomized experiment with individual noncompliance**) (Frangakis et al., 2002, Small et al., 2008). More details on statistical matching and inference in the context of preference-based IV analyses are discussed in detail in Zhang et al. [2020] and sketched in Section 6.

## 3 Eligibility and exclusion criteria

We consider the following eligibility and exclusion criteria that lead to the final study cohort. Number of subjects left in the study after applying each exclusion criterion is indicated in the parentheses.

1. Identify all non-duplicate beneficiaries undergoing isolated CABG, i.e. any case with an ICD-9 code for CABG surgery (n = 530,769);
2. Exclude beneficiaries in managed care (MC) and therefore without Part B claims (n = 406,588);
3. Exclude beneficiaries without at least six months of coverage prior to index admission (n = 380,041);
4. Exclude beneficiaries younger than 65 years of age (n = 344,009);
5. Exclude beneficiaries without a cardiac surgery related DRG code (n = 342,368);
6. Exclude years prior to 2013 (n = 117,519);
7. Exclude beneficiaries who had a stroke diagnosis within the six months prior to index admission for cardiac surgery and those who had a stroke that was “present on admission” (POA), and therefore not a complication post-cardiac surgery (n = 114,871);
8. When analyzing data at a surgeon-level granularity, we further exclude beneficiaries without a CPT code for CT surgery (n = 101,522).

TEE billing information is coded on CPT codes found in Part B claims. Step 2 excludes beneficiaries in managed care, who enrolled in fee-for-service, and therefore were without Part B, physician billing claims. Step 4 excludes beneficiaries enrolled in Medicare for disability or for end-stage renal disease. Step 5 excludes atypical beneficiaries of the procedure or had procedural miscoding.

## 4 Instrumental variable and IV-outcome confouders

We propose to leverage *hospitals’ (or surgeons’) preference* as an instrumental variable for patients’ receiving TEE monitoring during CABG surgery. We calculate the fraction of TEE usage out of CABG surgeries in our cohort for each hospital (or surgeon), and use this fraction as a measure of hospitals’ (or surgeons’) preference. Hospitals’ (or surgeons’) preference is identical for all patients treated in the same hospital (or by the same surgeon) and thus is a *cluster-level* IV. Importantly, although the IV is identical for different units within each cluster, there is *individual noncompliance*, i.e., each individual patient may or may not actually receive TEE monitoring during his/her CABG surgery.

An ideal instrumental variable has its variation completely exogenous; for instance, this is the case if draft lottery is used as an IV. In the context of observational studies, the results of an IV analysis may be biased if the IV and outcome are related by the so-called “instrument-outcome confounders” (Garabedian et al., 2014). In particular, Garabedian et al. [2014] pointed out the following potential instrument-outcome confounders: patients’ race, socioeconomic status, clinical risk factors, health status, facility and procedure volume, etc. Hospitals’ (or surgeons’) preference is more likely to be a valid instrumental variable after controlling for these IV-outcome confounding variables. We follow advice in Garabedian et al. [2014] and control for these IV-outcome confounding variables in our study.

Specifically, we control for two types of IV-outcome confounding variables: 1) unit-level confounding variables and 2) cluster-level confounding variables. Unit-level confounding variables refer to those related to each patient in the cluster and include the following:

- age;
- gender;
- race (white or not);
- nature of the surgery (elective or not),

and the following seven comorbid conditions:

- arrhythmia;
- congestive heart failure;
- diabetes;
- hypertension;
- obesity;
- pulmonary diseases;
- renal diseases.

We will further assume that hospitals’ (or surgeons’) preference and patients’ clinical outcomes are confounded by above confounding variables via some aggregate measures, e.g., the average age of patients in the cluster, percentage of patients who are male, percentage of patients with arrhythmia, etc.

Cluster-level confounding variables refer to characteristics related to each hospital (or surgeon), and include the following:

- total number of hospital beds;
- total number of full-time registered nurses;
- whether or not the hospital is a teaching hospital;
- whether or not the hospital is equipped with a cardiac intensive care unit;
- total cardiac surgery volume between January 1st, 2013 and October 15th, 2015 of the hospital (or the surgeon).

These cluster-level covariates are obtained from the American Hospital Association and merged to the patient-level Medicare and Medicaid claims data using organization/surgeon identifier numbers.

Left panel of Figure 1 plots the standardized differences for these covariates when comparing hospitals with IV (TEE usage preference) above the median and those below the median. Right panel of Figure 1 plots the same information at the surgeon level. Figure 1a suggests that hospitals that have a strong preference for TEE usage tend to have more hospital beds, more full-time registered nurses, larger surgical volume, and are more likely to be teaching hospitals. Patients in hospitals with larger preference for TEE usage also tend to be older, and more often have preexisting comorbid conditions including arrhythmia, pulmonary, and renal diseases. We saw a similar trend at the surgeon level.

**Figure 1:**
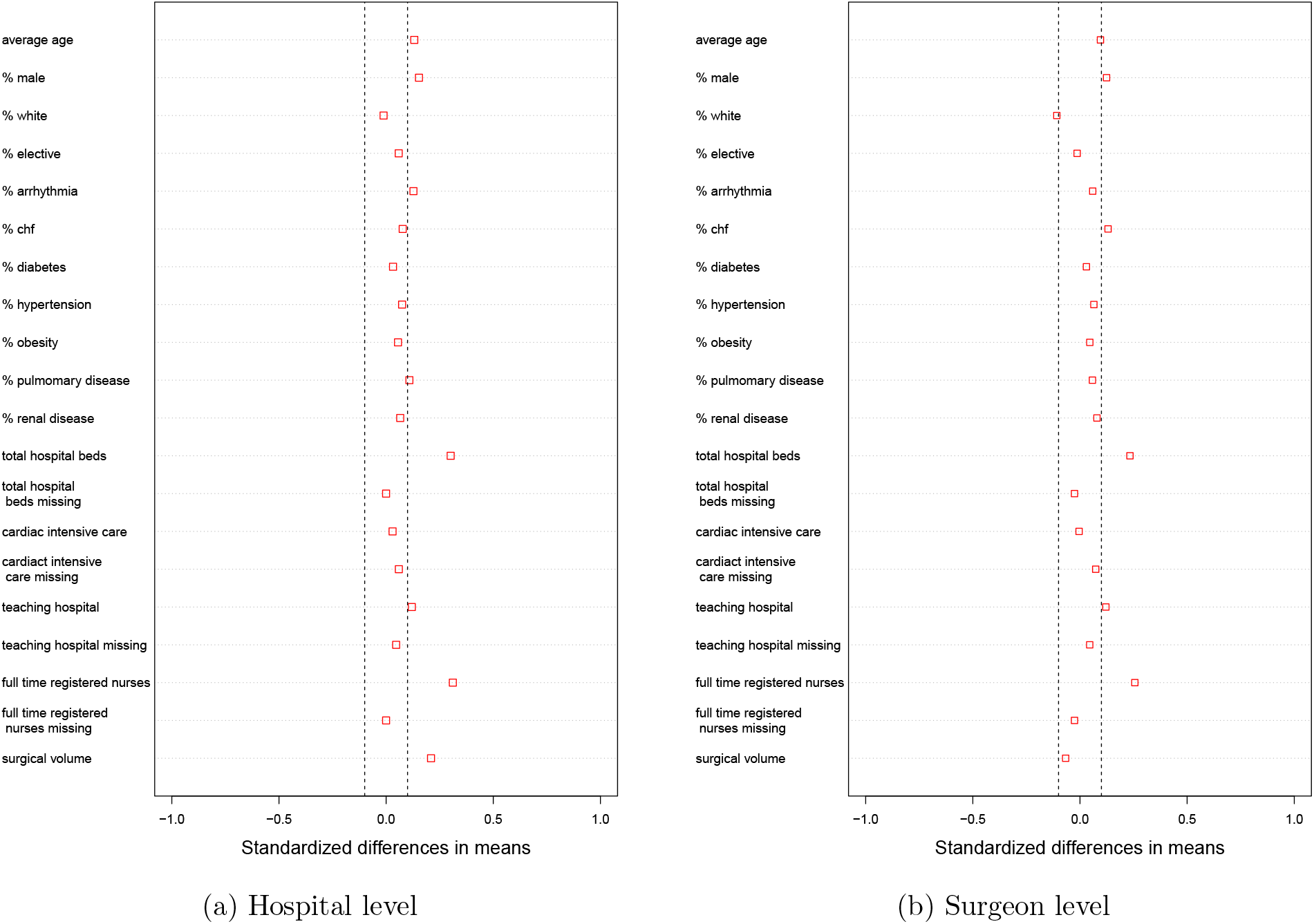
Love plot of standardized differences before matching. The standardized difference is calculated as (covariate mean of hospitals with IV above the median - covariate mean of hospitals with IV below the median) divided by the pooled within group standard deviation. Dashed lines are at -0.1 and 0.1 standardized differences.

## 5 Data granularity and outcomes

### 5.1 Data granularity

We will match the data at two different granularities: 1) hospital level and 2) surgeon level. Each hospital (or surgeon) defines a natural cluster of patients, i.e., those treated in the hospital or by the surgeon. We identified which patients belong to the same cluster (hospital or surgeon) using National Provider Identifier (NPI) numbers available in the CMS claims data. We will evaluate the quality of the match, including covariate balance, sample size, and difference in the encouragement dosage, and determine a granularity at which the primary analysis is carried out. The other granularity will be analyzed in a sensitivity analysis.

### 5.2 Primary outcomes

We consider one primary outcome: 30-day mortality.

### 5.3 Secondary outcomes

We consider three secondary outcomes:

1. 30-day mortality or incidence of stroke;
2. Length of hospitalization;
3. Incidence of esophageal perforation.

## 6 A matching-design approach to preference-based IV

### 6.1 Embedding observational IV data into a cluster-randomized experiment

Consider analyzing data at the hospital level. Ideally, we would like to pair two hospitals with identical patient-level and hospital-level confounding variables, but different preference for using TEE during CABG surgery. For instance, we would like to pair two hospitals, HA and HB, if HA and HB have exactly the same number of patients and identical composition of patients, e.g., each with two White males in their 80^;^s with three kinds of underlying comorbid conditions, etc. The hospital with a larger IV (hospital-level preference for TEE) will be considered the “encouraged” hospital and the other the “control” hospital.

However, such a strategy is very difficult if not impossible to implement. Instead, we consider the following assumption as described in (Zhang et al., 2020): hospitals’ (or surgeons’) preference is effectively randomized conditional on cluster-level covariates and some *aggregate measures* (e.g., mean and quantiles) of unit-level covariates within the cluster. In other words, the assumption says that it suffices to pair two hospitals (or surgeons) with the same (or at least very similar) average patient age, percentage of male patients, percentage of each important comorbid condition, etc, and cluster-level confounding variables like total number of hospital beds, total number of registered nurses, etc.

In practice, matching two hospitals (or surgeons) with identical percentage of male patients and identical percentage of each comorbidity is still very difficult. Instead in practice, researchers typically aim to create matched pairs in such a way that the mean values of each covariate in two groups after matching, i.e., the “encouraged” group and the “control” group, are as similar as possible. This is known as the covariate balance and can be assessed after each attempt to produce a match, without looking at the outcome data at all. We will regard a match as acceptable only if the standardized differences of all covariates are less than 0.1. We will also test if there is a statistically significant difference for each covariate in two groups after matching using a two-sample t-test. We will report the p-values and ideally only approximately 1 out of 20 p-values should be smaller than 0.05. If it turns out to be impossible to achieve this described covariate balance, we give priority to creating two well-comparable groups, possibly by discarding certain clusters, in order to ensure the internal validity of the study. When multiple matches could satisfy the desired covariate balance criteria, the match with a larger difference in the IV is preferred.

One way to achieve good covariate balance while maximizing the difference in the preference-based IV is to minimizing the robust Mahalanobis distance while penalizing two hospitals (or surgeons) having a similar preference using a technique called non-bipartite matching (Lu et al., 2011), also known as the near-far matching in the IV context (Baiocchi et al., 2010, Baiocchi et al., 2012, Neuman et al., 2014). In practice, penalties are also added to certain covariates to enforce better balance on these covariates.

### 6.2 Dealing with missing data

When merging the patient-level Medicare and Medicaid claims data with the cluster-level American Hospital Association data, we sometimes encounter missing data. This happens when we cannot identify certain patients’ hospital and when AHA data contains missing data on cluster-level covariates for certain hospitals. We will deal with missing data by creating one “missing data indicator” covariate for each covariate with missing data, and balance these missing data indicators in the encouraged and control groups.

### 6.3 Software

We will be performing the described statistical matching using the R package `nbpMatching` (Lu et al., 2011, Beck et al., 2016) and summarizing the covariate balancing using the R package `RItools` (Hansen and Bowers, 2008, Bowers et al., 2019).

## 7 Inference

Suppose that there are *K* matched pairs of two clusters (hospitals or surgeons). Suppose that cluster *j* in matched pair *i* contains *n_kj_* units (i.e., patients). Let 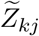 denote the observed IV of cluster *k* in matched pair *j*, *k* = 1,…,*K* and *j* = 1,2. For each *k*, let 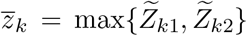 denote the observed IV for the encouraged cluster in matched pair *k*, and 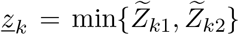 for the control cluster in matched pair *k*. Let 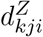 and 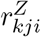 denote the potential treatment indicator (using TEE or not) and the potential outcome that we want to study (e.g., can be either the primary or secondary outcome) of unit *i* in cluster *j* in matched pair *k* given that the unit’s IV score is *Z* respectively, where *k* = 1,…, *K*, *j* = 1,2, and *i* = 1…, *n_kj_*. Let *D_kji_* and *R_kji_* denote the observed treatment indicator and the observed outcome of unit *i* in cluster *j* in matched pair *k* respectively, where *k* = 1, …, *K*, *j* = 1,2, and *i* = 1 …,*n_kj_*. Following Small et al. [2008] and Zhang et al. [2020], we consider the following cluster-level proportional treatment effect model:

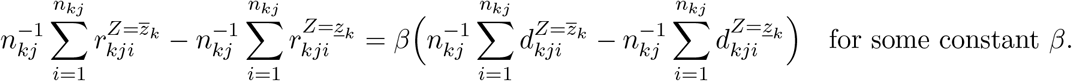

We focus on testing 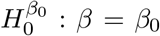 versus 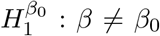 for various *β*_0_ (perhaps focusing on *β*_0_ = 0) and build a two-sided 95% confidence interval for *β*. We will test 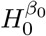 versus 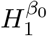 by using the `senm` command in `R` package `sensitivitymult` (Rosenbaum, 2017), which performs the paired Huber’s M test in a randomization inference (Rosenbaum, 2007), with the input vector of responses 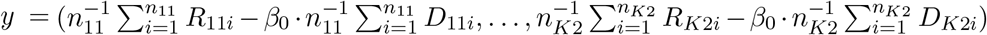. When using the `senm` command, we set all parameters to their default values (i.e., `gamma` = 1, `inner` = 0, `trim` = 3, `lambda` = 1/2, `tau` = 0, `alternative`=“greater”, `TonT`= FALSE), and get the corresponding deviate. We then take the square of the output deviate and then get the two-sided p-value by comparing the squared deviate with the quantile function of the chi-square distribution with one degree of freedom. We will report a two-sided *p*-value using the above procedure for the null hypothesis *H*_0_: *β* = 0 versus *H*_1_: *β* ≠ 0 (i.e., 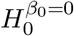 versus 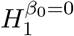). We will also report a 95% confidence interval for *β* (found by inverting the hypothesis test for 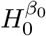 under various *β*_0_ using the above procedure; see Rosenbaum, 2002). We conduct this statistical analysis procedure for all the primary and secondary outcomes.

## 8 Sensitivity analysis

We follow the framework in Rosenbaum [1989] to conduct a sensitivity analysis for the conclusion drawn from the primary analysis. To be specific, we assume that IV dose assignment probability within the *k*-th matched pair can be biased by

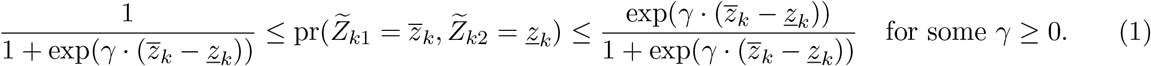

To conduct a two-sided *α*-level sensitivity analysis, we follow the procedure described in Rosenbaum [1989] to perform two one-sided *α/2* sensitivity analyses for testing *H*_0_: *β* = 0 with the paired Huber’s M test statistic used in Section 7 under the constraint (1) and various sensitivity parameters γ. For each γ ≥ 0, we reject the null hypothesis of no treatment effect *H*_0_: *β* = 0 if and only if we reject *H*_0_: *β* = 0 in either one of the two one-sided *α*/2-level sensitivity analyses under the sensitivity parameter γ, forming a two-sided *α*-level sensitivity analysis. We gradually increase γ until we fail to reject and denote such changepoint γ as 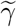. We then report the average magnitude of the biased ratios of the odds of receiving encouragement among the *K* matched pairs (i.e., 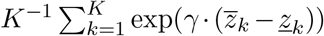) to alter the conclusion drawn from the primary analysis under level *α* = 0.05.

## 9 Match Results

### 9.1 Hospital-level match

Table 2 summarizes a hospital-level match that meets the prespecified covariate balance criteria. There are a total of 1,215 hospitals that have performed at least one CABG surgery subject to the eligibility and exclusion criteria described in Section 3. We formed 484 matched pairs of 2 hospitals using 968 out of 1,215 hospitals (79.7%) without ties on the IV. These 968 hospitals contain a total of 91, 516 patients, accounting for 79.7% of all patients in our study population. Table 2 suggests that we have achieved very good covariate balance, with all standardized differences less than 0.1. We also perform a simple two-sample t-test for each confounding variable and no confounding variable is significant at 0.1 level. For a graphical contrast of before and after matching covariate balance, see Figure 2a. Moreover, hospitals’ preference in the encouraged group is still remarkably different from that in the control group (0.37 vs. 0.72).

**Table 1:** Covariate balance **before matching**. We compare hospitals (or surgeons) with TEE preference above the median to those with TEE preference below the median. Significance codes: “*” 0.05; “**” 0 01 “***” 0 001

|  | Hospital level |  |  | Surgeon level |  |  |
| --- | --- | --- | --- | --- | --- | --- |
|  | Below Median | Above Median | Standardized Difference | Below Median | Above Median | Standardized Difference |
| Instrumental variable: proportion of TEE used in CABG surgeries | 0.27 | 0.81 | 3.61 | 0.39 | 0.95 | 3.09 |
| **Patient-level Characteristics** |  |  |  |  |  |  |
| Average age | 75.10 | 75.33 | 0.13* | 75.25 | 75.53 | 0.10*** |
| % male | 0.67 | 0.69 | 0.15** | 0.68 | 0.70 | 0.12*** |
| % white | 0.85 | 0.85 | -0.01 | 0.88 | 0.86 | -0.11*** |
| % elective | 0.46 | 0.47 | 0.06 | 0.48 | 0.47 | -0.01 |
| % with arrhythmia | 0.10 | 0.11 | 0.13* | 0.11 | 0.12 | 0.06* |
| % with congestive heart failure | 0.11 | 0.11 | 0.08 | 0.10 | 0.12 | 0.13*** |
| % with diabetes | 0.16 | 0.16 | 0.03 | 0.16 | 0.16 | 0.03 |
| % with hypertension | 0.27 | 0.28 | 0.07 | 0.28 | 0.30 | 0.06* |
| % with obesity | 0.05 | 0.06 | 0.06 | 0.06 | 0.06 | 0.05 |
| % with pulmonary diseases | 0.02 | 0.02 | 0.11 | 0.02 | 0.02 | 0.06* |
| % with renal diseases | 0.08 | 0.09 | 0.07 | 0.08 | 0.10 | 0.08** |
| **Cluster-level Characteristics** |  |  |  |  |  |  |
| Hospital beds | 340.79 | 415.81 | 0.30*** | 449.83 | 521.65 | 0.23*** |
| Hospital beds (missing) | 0.01 | 0.01 | 0.00 | 0.01 | 0.01 | -0.03 |
| Cardiac ICU (yes/no) | 0.71 | 0.72 | 0.03 | 0.83 | 0.83 | 0.00 |
| Cardiac ICU (missing) | 0.07 | 0.09 | 0.06 | 0.04 | 0.05 | 0.07** |
| Teaching hospital | 0.15 | 0.20 | 0.12* | 0.27 | 0.33 | 0.12*** |
| Teaching hospital (missing) | 0.00 | 0.01 | 0.05 | 0.00 | 0.00 | 0.05 |
| Full-time nurses | 539.33 | 717.86 | 0.31*** | 798.56 | 1001.79 | 0.26*** |
| Full-time nurses (missing) | 0.01 | 0.01 | 0.00 | 0.01 | 0.01 | -0.03 |
| Total surgical volume | 456.74 | 572.67 | 0.21*** | 68.96 | 63.87 | -0.07* |

**Table 2:**
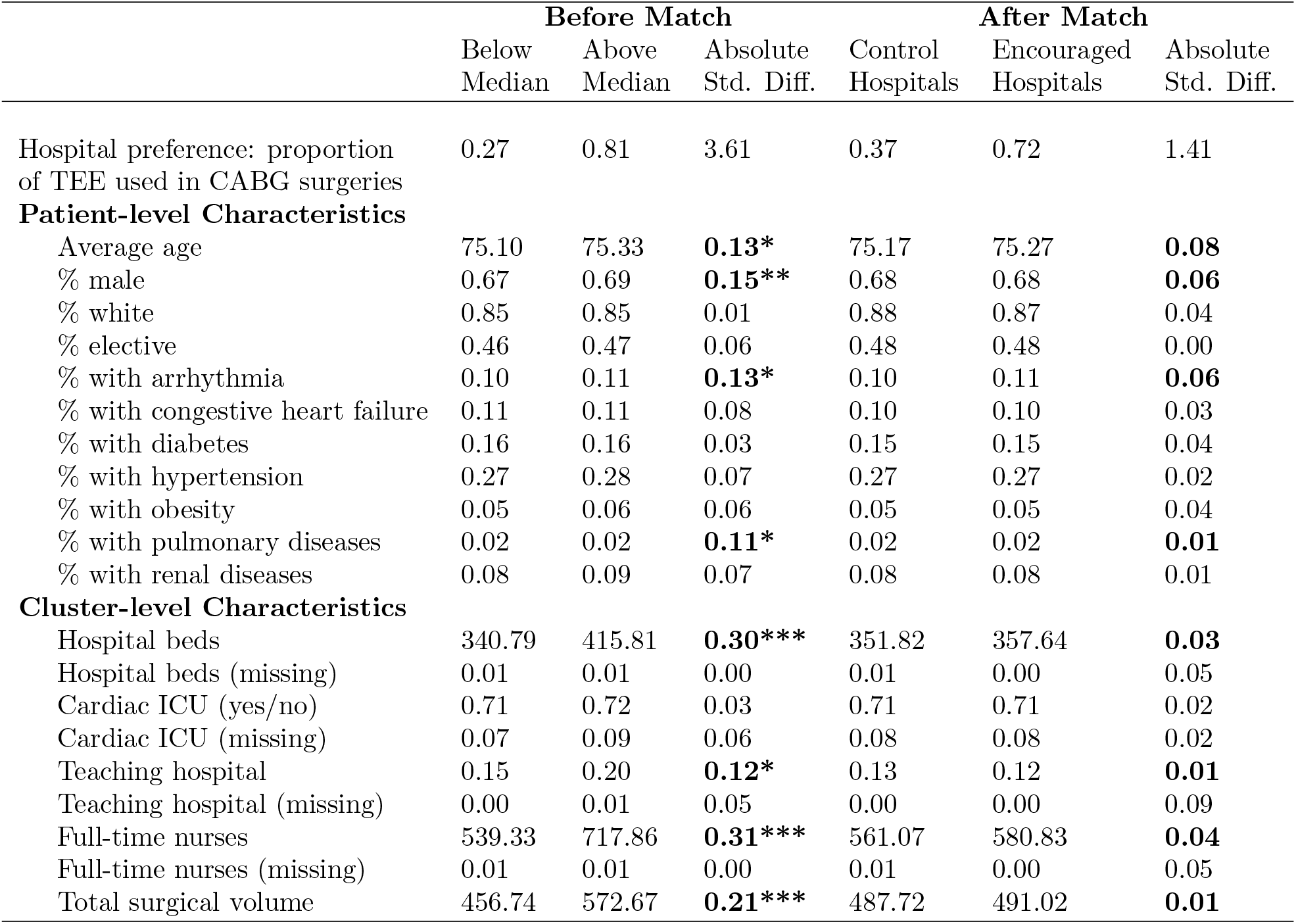
Covariate balance at **hospital level** before and after matching. **Before matching:** these three columns are duplicated from Table 1 and included here for convenience of comparison. **After matching:** 484 pairs of 2 hospitals are formed after matching. These 968 hospitals represent a total of 91, 516 patients. Matching helps largely improve the covariate balance for a few key variables (highlighted). After matching, no covariate is statistically different in two groups at 0.1 level. Significance codes: “*” 0.05; “**” 0.01; “***” 0.001.

**Figure 2:**
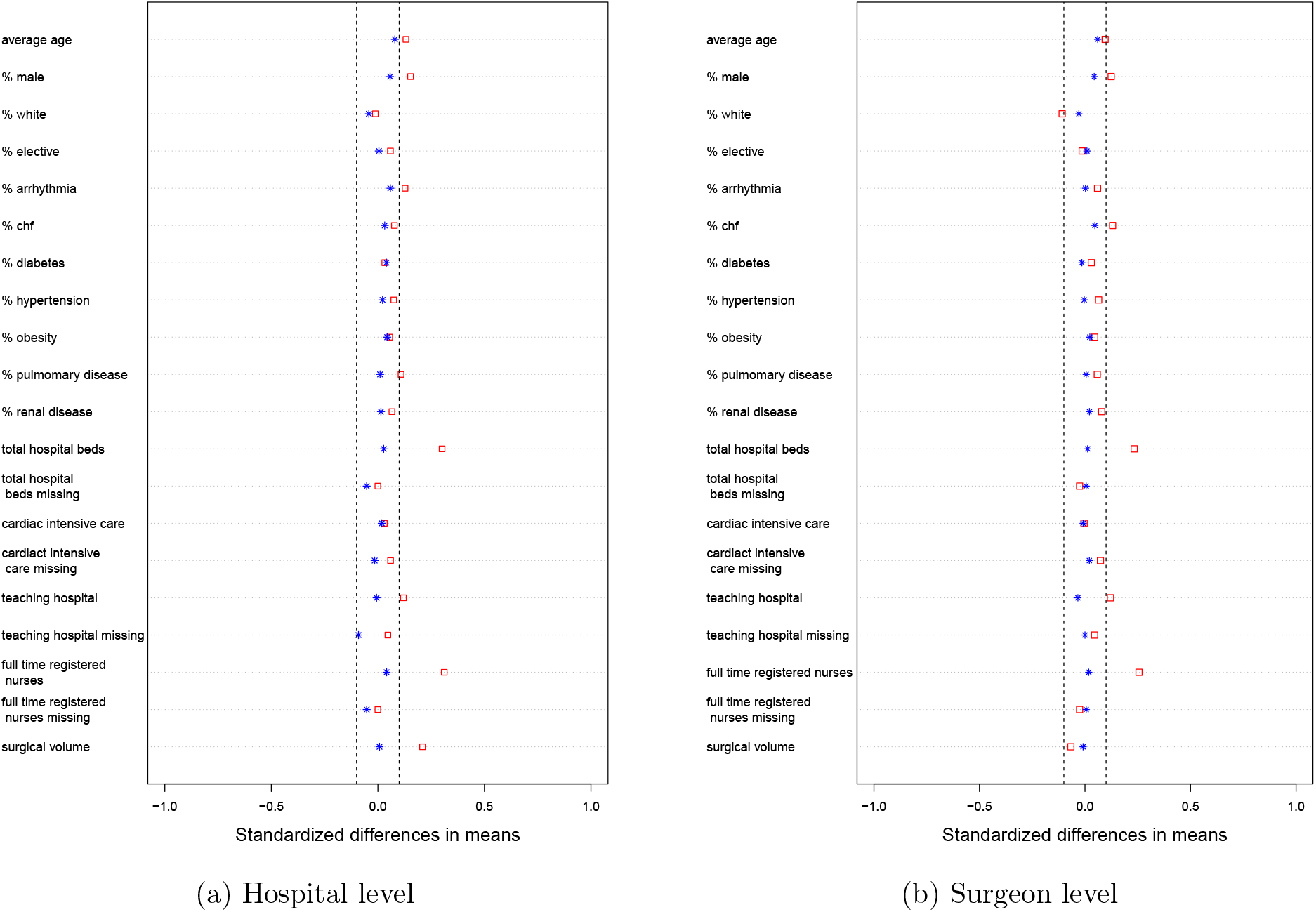
Love plot of standardized differences before (red squares) and after (blue asterisks) matching. Standardized difference before matching is calculated as (covariate mean of counties above the median preference for TEE - covariate mean of counties below the median preference for TEE) divided by the pooled within group standard deviation. Dashed lines are at -0.1 and 0.1 standardized differences.

### 9.2 Surgeon-level match

Similarly, at the surgeon level, we are able to identify a total of 5, 012 surgeons who have performed at least one CABG surgery on our study cohort. We are able to form 2,113 pairs of two surgeons using 4, 226 out of all 5, 012 (84.3%) surgeons without ties on the IV. These 4, 226 surgeons represent a total of 95, 275 patients, accounting for 93.8% of the total study cohort available at the surgeon level. Table 3 summarizes the covariate balance and Figure 2b gives a graphical representation. Again, the covariate balance is stellar: all standardized differences are well below 0.1 and no covariate is statistically different at 0.05 level.

**Table 3:** Covariate balance at **surgeon level** before and after matching. **Before matching:** these three columns are duplicated from Table 1 and included here for convenience of comparison. **After matching:** 2113 pairs of 2 surgeons are formed after matching. These 4, 226 hospitals represent a total of 95, 275 patients. Matching helps largely improve the covariate balance for a few key variables (highlighted). After matching, no covariate is statistically different in two groups at 0.05 level. Significance codes: “*” 0.05; “**” 0.01; “***” 0.001.

|  | Before Match |  |  | After Match |  |  |
| --- | --- | --- | --- | --- | --- | --- |
|  | Below<br>Median | Above<br>Median | Absolute<br>Std. Diff. | Control<br>Hospitals | Encouraged<br>Hospitals | Absolute<br>Std. Diff. |
| Surgeon preference: proportion<br>of TEE used in CABG surgeries | 0.39 | 0.95 | 3.09 | 0.44 | 0.85 | 1.50 |
| **Patient-level Characteristics** |  |  |  |  |  |  |
| Average age | 75.25 | 75.53 | 0.10*** | 75.28 | 75.43 | 0.06 |
| % male | 0.68 | 0.70 | **0.12\*\*\*** | 0.68 | 0.69 | **0.04** |
| % white | 0.88 | 0.86 | **0.11\*\*\*** | 0.88 | 0.88 | **0.03** |
| % elective | 0.48 | 0.47 | 0.01 | 0.47 | 0.47 | 0.01 |
| % with arrhythmia | 0.11 | 0.12 | 0.06* | 0.11 | 0.11 | 0.00 |
| % with congestive heart failure | 0.10 | 0.12 | **0.13\*\*\*** | 0.10 | 0.10 | **0.05** |
| % with diabetes | 0.16 | 0.16 | 0.03 | 0.15 | 0.15 | 0.01 |
| % with hypertension | 0.28 | 0.30 | 0.06* | 0.28 | 0.28 | 0.00 |
| % with obesity | 0.06 | 0.06 | 0.05 | 0.05 | 0.06 | 0.02 |
| % with pulmonary diseases | 0.02 | 0.02 | 0.06* | 0.02 | 0.02 | 0.01 |
| % with renal diseases | 0.08 | 0.10 | 0.08** | 0.08 | 0.08 | 0.02 |
| **Cluster-level Characteristics** |  |  |  |  |  |  |
| Hospital beds | 449.83 | 521.65 | **0.23\*\*\*** | 450.34 | 453.68 | **0.01** |
| Hospital beds (missing) | 0.01 | 0.01 | 0.03 | 0.01 | 0.01 | 0.01 |
| Cardiac ICU (yes/no) | 0.83 | 0.83 | 0.00 | 0.83 | 0.83 | 0.01 |
| Cardiac ICU (missing) | 0.04 | 0.05 | 0.07** | 0.04 | 0.05 | 0.02 |
| Teaching hospital | 0.27 | 0.33 | **0.12\*\*\*** | 0.26 | 0.25 | **0.03** |
| Teaching hospital (missing) | 0.00 | 0.00 | 0.05 | 0.00 | 0.00 | 0.00 |
| Full-time nurses | 798.56 | 1001.79 | **0.26\*\*\*** | 800.23 | 812.10 | **0.02** |
| Full-time nurses (missing) | 0.01 | 0.01 | 0.03 | 0.01 | 0.01 | 0.01 |
| Total surgical volume | 68.96 | 63.87 | 0.07* | 69.40 | 68.73 | 0.01 |

### 9.3 Deciding a data granularity for primary analysis

Both the hospital-level and the surgeon-level matches have achieved good balance, and the difference in the IV dosage is similar. Although we have paired two surgeons who have very similar cardiac surgical volume and work in very similar hospitals, there might still remain some residual confounding that biases surgeons’ preference and affect the clinical outcomes, e.g., surgeons’ education. Therefore, we choose to analyze the data at the hospital level with the hospital-level match, and leave the surgeon-level analysis to the sensitivity analysis.

## 10 Sensitivity analyses

We will analyze the data at the surgeon level using the surgeon-level match as a sensitivity analysis.

## Data Availability

All data used for this study are available for purchase from the Centers for Medicare and Medicaid Services (CMS). This work was conducted in accordance with a Data Use Agreement (DUA) with CMS. We are not permitted to share the data directly.

